# Regional cerebral atrophy contributes to personalized survival prediction in ALS: a multicentre, machine learning, deformation based morphometry study

**DOI:** 10.1101/2024.10.04.24314899

**Authors:** Isabelle Lajoie, Canadian ALS Neuroimaging Consortium (CALSNIC), Sanjay Kalra, Mahsa Dadar

## Abstract

**Objective:** Accurate personalized survival prediction in amyotrophic lateral sclerosis is essential for effective patient care planning. This study investigates whether gray and white matter changes measured by magnetic resonance imaging can improve individual survival predictions.

**Methods:** We analyzed data from 178 amyotrophic lateral sclerosis patients and 166 healthy controls in the Canadian ALS Neuroimaging Consortium study. A voxel-wise linear mixed-effects model assessed disease-related and survival-related atrophy detected through deformation-based morphometry, controlling for age, sex, and scanner variations. Additional linear mixed-effects models explored associations between regional imaging and clinical measurements, and their associations with time to the composite outcome of death, tracheostomy or permanent assisted ventilation. An individual survival distributions model was evaluated using clinical data alone, imaging data alone, and a combination of both features.

**Results:** Deformation-based morphometry uncovered distinct voxel-wise atrophy patterns linked to disease progression and survival, with many of these regional atrophy significantly associated with clinical manifestations of the disease. By integrating regional imaging features with clinical data, we observed a substantial enhancement in the performance of survival models across key metrics. Our analysis identified specific brain regions, such as the corpus callosum, rostral middle frontal gyrus, and thalamus, where atrophy predicted an increased risk of mortality.

**Interpretation:** This study suggests that brain atrophy patterns measured by deformation-based morphometry provide valuable insights beyond clinical assessments for prognosis. It offers a more comprehensive approach to prognosis and highlights brain regions involved in disease progression and survival, potentially leading to a better understanding of amyotrophic lateral sclerosis.

## Introduction

Amyotrophic lateral sclerosis (ALS) presents with a wide range of clinical manifestations, complicating accurate prognosis. Factors contributing to this clinical heterogeneity include age of onset, mix of Upper Motor Neuron (UMN) and Lower Motor Neuron (LMN) signs, disease duration, cognitive and behavioral changes, and comorbid conditions^1^. Post-diagnosis, mortality and survival duration vary significantly, typically between two to five years, with 5-10% of ALS patients surviving longer than 10 years^2^. This variability likely results from the coexistence of different pathogenic mechanisms that are challenging to quantify and disentangle^3^.

The severity and patterns of brain atrophy also vary widely among ALS patients^4^, with both grey matter (GM) and white matter (WM) showing varying degrees of atrophy in primarily motor but also extra-motor areas^4^. This clinical and biological heterogeneity may partly explain the absence of an effective treatment^5^. Understanding the relationship between brain atrophy patterns and the heterogeneous clinical features of the disease could lead to more defined characterization of the patients’ pathological profiles, leading to the development of focused personalized therapeutics.

MRI offers promise in developing biomarkers for quantification of brain atrophy and disease progression in ALS. It allows for non-invasive regional brain assessments with minimal risk and is objective, widely available, and repeatable. Deformation-based morphometry (DBM) is a sensitive and reliable technique for the quantification of GM and WM atrophy using T1- weighted structural MRIs^6^. Previous DBM-based studies have detected progressive cerebral atrophy in various neurodegenerative disorders^7–10^.

MRI imaging features can also be integrated into machine learning models to predict survival outcomes^11–15^. However, existing limitations have hindered the development of robust survival models in ALS, primarily due to small sample sizes or single-center cohorts^12–14^, or a unique focus on gray matter GM changes^11^ that overlooks the potential contribution of WM integrity to disease progression. This is particularly important given the significant neurodegeneration of WM areas such as the corticospinal tract (CST) and corpus callosum, in ALS patients^12,16,17^. Additionally, many studies approach the survival prediction task as a binary classification for a specific single-time point^14,18,19^, limiting personalized prognoses and introducing bias^20,21^ by excluding censored patients, such as those lost to follow-up or seeking medical assistance. In contrast to single-time point classification, individualized survival distribution (ISD) models effectively incorporate censored patients and offer more refined and personalized prognosis by estimating the probability of patient survival at different future time points.

The objective of this study was to determine the association between brain atrophy patterns and individual survival in ALS patients by leveraging the CALSNIC dataset, a comprehensive, large-scale collection of deeply phenotyped data critical for our analysis. We utilized DBM to evaluate cross-sectional and longitudinal brain changes in ALS, focusing on how these changes correlate with shorter survival times and clinical outcomes. Machine learning techniques, with a focus on ISD models, were employed to evaluate whether integration of imaging features enhances the predictive accuracy of individualized survival probabilities. Ultimately, this approach aims to deliver more personalized prognostic insights for ALS patients, guiding treatment decisions and optimizing care.

## Method

### Participants

We used longitudinal (baseline, months 4 and 8) MRI and clinical measurements of 230 ALS patients and 204 healthy controls (HCs) from the CALSNIC study^22^. CALSNIC adheres to the principles of the Declaration of Helsinki and received approval from the Health Research Ethics Boards of all participating sites. Written informed consent was obtained from all participants and the University of Alberta Health Research Ethics Board (HREB) granted approval for the protocol presented in this study. Clinical and MRI protocols were harmonized across research centers and vendors. Following the exclusion criteria (detailed in the supplementary materials, Section.A), the final study population included 178 ALS patients (175_baseline_/107_visit2_/67_visit3_) and 166 HCs (165_baseline_/128_visit2_/98_visit3_) (Fig 1A).

**Figure 1.**
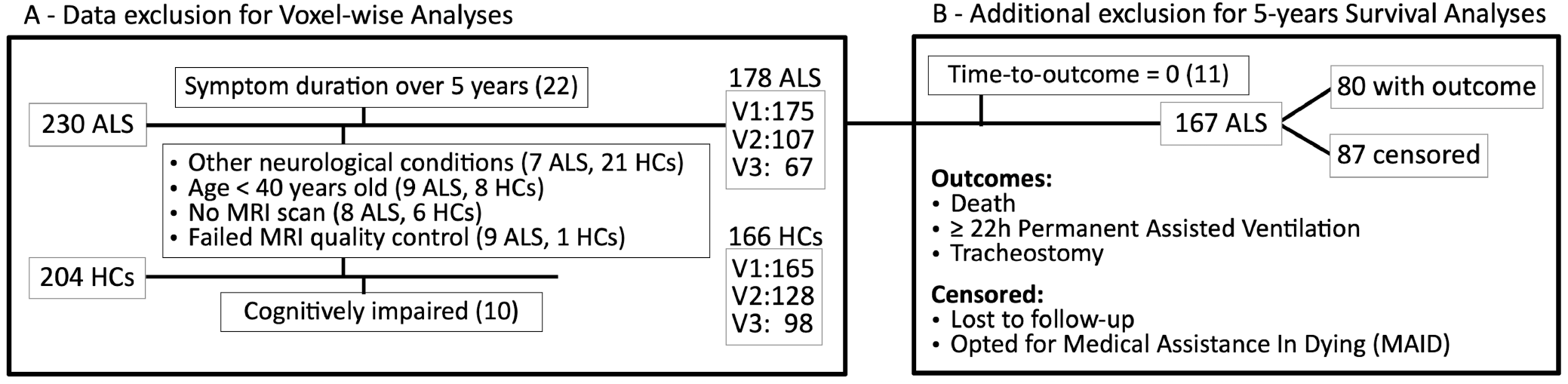
Participants exclusion: Participants exclusion flowchart for the voxel-wise **(A)** and survival analyses **(B)**. ALS: Amyotrophic lateral sclerosis patients; HCs: healthy controls cohorts; MRI: magnetic resonance imaging.

Survival outcome was defined as death, tracheostomy, or assisted ventilation for at least 22 hours. Patients were censored if they were alive five years after the baseline MRI visit, lost to follow-up, or if they opted for medical assistance in dying (MAID). Patients with no follow-up after the initial MRI visit were excluded. The final survival dataset included 167 patients, with a censoring rate of 52% (Fig 1B). Patients were further categorized as “short” or “long” survivors based on whether their time-to-outcome from the baseline MRI visit was before or after 24 months, respectively. Censored patients were excluded from this classification analysis if their censoring time occurred before 24 months. Clinical evaluation included the ALS Functional Rating Scale-Revised (ALSFRS-R)^24^, disease progression rate (DPR), finger and foot tapping rates, Edinburgh Cognitive and Behavioural ALS Screen (ECAS), UMN and LMN involvement, forced vital capacity (FVC), symptom duration at the first MRI visit, ethnicity, years of education, handedness, age and sex. For details on the clinical evaluations, see Section.B in the supplementary materials.

### MRI processing

All T1-weighted MRIs were processed using our open source and extensively validated pipeline (https://github.com/VANDAlab/Preprocessing_Pipeline) based on the open access MINC (https://github.com/BIC-MNI/minc-tools) and ANTs (http://stnava.github.io/ANTs/) tools. Pre-processing steps included: denoising^27^, intensity inhomogeneity correction^28^, and intensity normalization using histogram matching. The images were linearly^29^ and nonlinearly^30^ registered to the MNI-ICBM152-2009c template^31^. All steps were visually assessed to exclude cases with significant artifacts or inaccurate registrations. DBM maps were calculated as the Jacobian determinant of the non-linear deformation fields. DBM values greater than one indicate localized expansion compared to the corresponding voxels in the template, whereas values smaller than one indicate relative shrinkage, i.e. atrophy. For regional analyses, DBM maps were masked to remove the partial volume effects of the sulci and mean DBM was calculated based on 62 bilateral GM and ventricle regions from the CerebrA atlas^34^, and WM tracts and corpus callosum based on the JHU^35^ and Allen^36^ atlases, respectively.

### Statistical analyses

Clinical and demographic measures were compared between patients and controls, as well as short and long survivors. Normality was established employing the Shapiro-Wilk test. Continuous variables were compared employing two-tailed *t*-tests and Mann-Whitney U tests, while categorical variables were compared using Chi-squared tests. A *p*-value threshold <0.05 was employed for statistical significance. For each metric, participants with missing value were excluded from the computed statistics.

### Brain atrophy

To investigate the disease-related brain changes, the following voxel-wise mixed-effects (LME) model was applied:

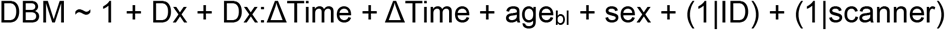

where DBM indicates the voxel-wise DBM value. Dx is a categorical fixed variable contrasting ALS vs HCs to evaluate cross-sectional brain changes. Dx:ΔTime denotes the interaction between diagnostic group and follow-up time, reflecting the difference in the rate (slope) of the longitudinal brain changes between the patients and controls. Age_bl_ indicates the participant’s age at the time of the first visit and sex is a categorical variable contrasting male vs female. Participant ID and scanner site were included as categorical random variables. An additional LME model was employed to investigate voxel-wise differences between survival groups and controls. Dx indicated ALS short and long survival groups and HCs. Finally, a third LME model was applied to specifically compare the short and long survival groups, to determine the regions associated with survival. Results were corrected for multiple comparisons using the Benjamini and Hochberg/Yekutieli false discovery rate (FDR) controlling technique^32,33^ with a statistical significance threshold of 0.05. LME analyses were performed using ‘fitlme’ in MATLABv2021a.

### Association between brain atrophy and clinical symptoms

To examine the relationship between clinical and regional imaging measurements (DBM), we conducted LME analyses using the following model:

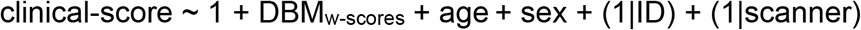

A second LME analysis was performed on the cross-sectional data to evaluate the univariate association between each measurement and time-to-event outcomes in uncensored patients:

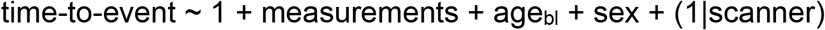

This approach provided insights into how clinical and neuroimaging factors influence survival. Results were corrected for multiple comparisons using FDR with a threshold of 0.05.

### Survival prediction

To focus on disease-specific atrophy in ALS patients, we calculated regional w-scores for each patient^37^. The w-score is the adjusted difference between a patient’s observed DBM value and its predicted value normalized for age, sex, and imaging site. This predicted value is derived from a linear mixed-effects model based on data from HCs. A negative w-score indicates greater atrophy compared to expected levels in the HCs. Clinical features with missing data in over 20% of the patients were excluded, yielding a final number of 83 clinical and demographic features.

To assess the contribution of DBM features to prognostic accuracy, we trained a Cox’s Proportional Hazards machine learning survival model with elastic-net penalty (CPH) using regional imaging (DBM w-scores) features alone, clinical features alone, and clinical features combined with imaging features. The model effectively handles censored participants and generates Individual Survival Distributions (ISD) for unseen patient data, offering detailed survival probabilities across all future time points. Nested cross-validation was employed to optimize the hyperparameters (3 folds) and to evaluate each model’s performance (5 folds), and was repeated 100 times with different random seeds for train/test stratification to ensure robust performance evaluation. The model’s discrimination capability was evaluated using the concordance index (C-index)^38^. Calibration, indicating how closely predicted survival probabilities match actual outcomes, was evaluated using the Integrated Brier Score (IBS)^39^ and D-calibration, also known as Distributional Calibration^40^, with a *p*-value greater than 0.05 indicating that the survival curves are well-calibrated. Finally, two variants of mean absolute error (MAE) that account for censored participants were computed based on predicted median survival times (50% survival probability): MAE-Margin^40^, which assigns an estimated value (margin time) to each censored subject using the non-parametric Kaplan-Meier estimator^41^, and MAE-PO, which uses pseudo-observation (PO) values as estimates^42^. Feature preprocessing was performed within the outer loop and involved imputing missing values (using the mean for numerical features and the most frequent value for categorical features), followed by robust scaling for numerical features and one-hot encoding for unordered categorical features. Feature preselection was performed in the outer loop, retrieving the k-best predictors from univariate Cox proportional models.

In addition to evaluating the models’ ability to predict individual survival probabilities across all future time points, we assessed their performance in predicting outcomes at a specific time- point using the ISDs. In this context of a binary classification task, adjusting the criterion threshold for event time prediction enables the optimization of sensitivity and specificity profiles according to specific requirements. We used the Youden index across averaged iterations while prioritizing sensitivity to enhance the model’s accuracy in correctly identifying ALS patients who experienced the event before the specified time point. This approach ensures that patients at higher risk of rapid progression can benefit from more aggressive therapeutic strategies and closer monitoring. Patients with censoring time preceding the target time-point were excluded from this analysis, as their status at that time could not be observed.

Our survival model measures each feature’s impact on the risk of the outcome using coefficients that are translated into hazard ratios (HR). Unlike univariate correlation analysis, these feature importances are derived from a multivariate machine learning model, accounting for the combined effect of all predictors. Feature importance was evaluated by averaging HR values across 100 iterations of nested cross-validation, assigning an HR of 1 (indicating no impact) to features not selected in specific iterations. We also computed feature importance using the entire dataset for comparison. We report HRs with 95% confidence intervals (CI), where an HR>1 indicates a higher risk of death or respiratory failure, while HR<1 indicates a higher risk with a decrease in the predictor. Survival analysis was performed using the Python-based packages scikit-learn^43^, scikit-survival^44^ and SurvivalEVAL^45^.

## Results

### Demographics

165 HCs and 176 patients were included, from which 101 had sufficient survival information to be classified into the short (41) or long (60) survival groups, considering their time-to-event being within 24 months or after, respectively. A summary of the participants’ demographics, cognitive, and functional performance is presented in Table 1. Mean age in the ALS patients was significantly greater than that of HCs. There was a greater proportion of men in ALS patients than HCs. Years of education were slightly lower in ALS patients compared to HCs. As expected, ALS patients showed greater cognitive and functional impairment. Significant differences were found in finger and foot tapping, and ECAS scores. There were no significant differences in age, sex and years of education between the long and short survival groups. Participants in the short survival group had higher DPR, lower FVC scores and higher LMN burden scores compared to the long survival group.

### Brain atrophy

The LME analyses revealed greater bilateral atrophy in the primary motor cortex, ventral diencephalon, brainstem, amygdala, thalamus and basal ganglia of the patients. Greater atrophy was also observed in major WM tracts including the CST, superior longitudinal fasciculus (posterior limb tracts), anterior thalamocortical tracts, and corpus callosum, accompanied by enlargement of the third ventricle in the ALS group (Fig 2A). Patients in the short survival group had greater atrophy and ventricular and sulcal expansion (Fig 2B) compared to the long survival group, whose atrophy was limited to the CST and medulla oblongata (Fig 2C). A direct comparison between ALS_short_ and ALS_long_ groups highlighted the more pronounced atrophy in the body and splenium of the corpus callosum in the shorter survival group.

**Figure 2.**
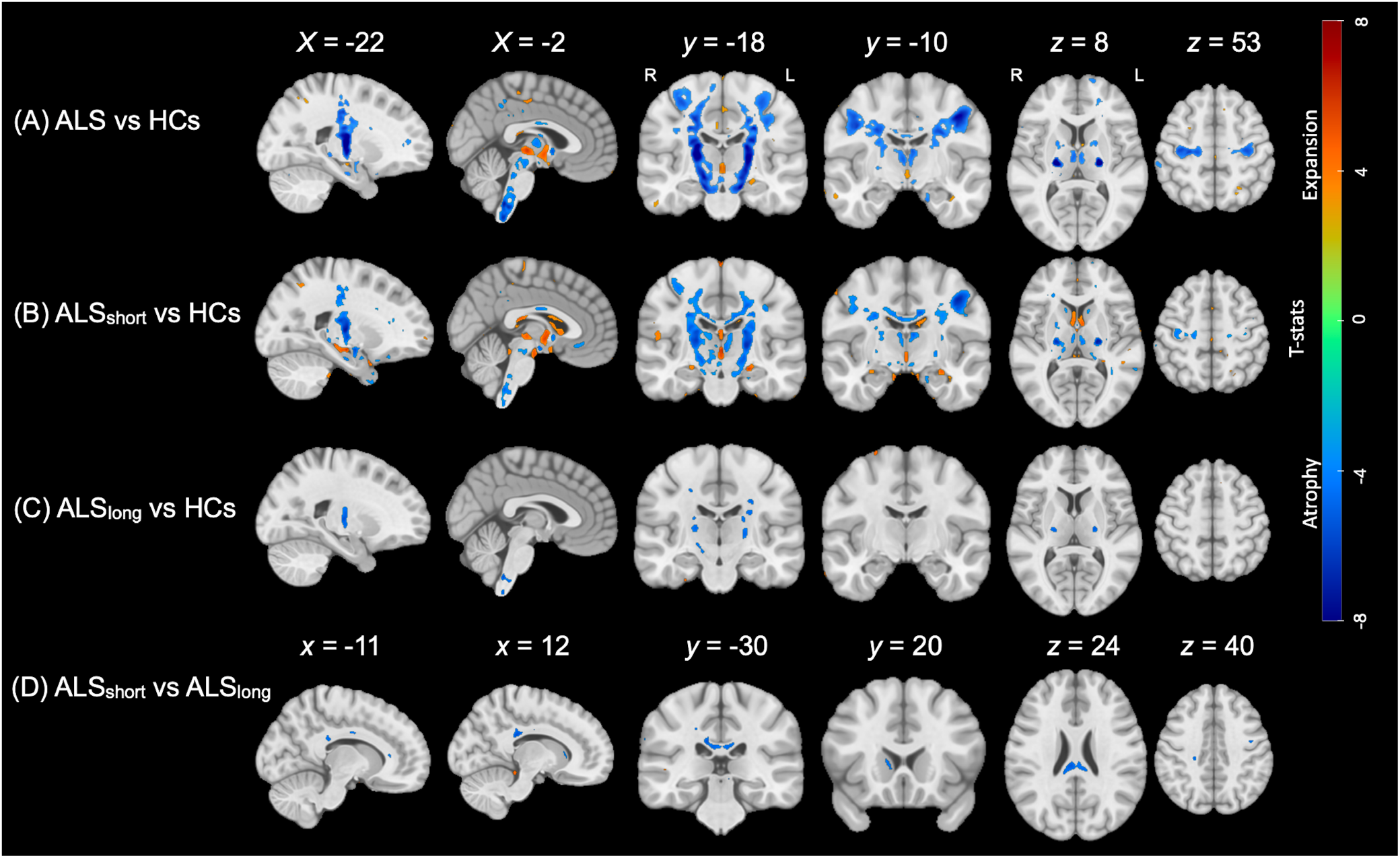
Cross-sectional Volumetric Changes: T-statistic maps (p-FDR < 0.05) illustrate cross-sectional differences (term Dx in the LME), overlaid on the MNI template. Panels display: ALS vs HCs **(A)** ALS_short_ vs HCs **(B)**, ALS_long_ vs HCs **(C)**, and ALS_short_ vs ALS_long_ **(D)**. ‘ALS_short_’ and ‘ALS_long_’ refer to patients who experienced the event prior to and after 24 months respectively. HCs represent healthy controls. Warmer colors highlight regions of enlargement (e.g., ventricular and sulcal regions), while cooler colors indicate areas of tissue atrophy.

The results further revealed additional progressive atrophy (indicated by the interaction term) in the primary motor cortex, caudate nucleus, and pars opercularis, the corpus callosum and superior longitudinal fasciculus, along with significant ventricular and sulcal enlargement. Changes in the full ALS group (Fig 3A) were mirrored in the ALS_long_ group (Fig 3C), with fewer regions detected in the ALS_short_ group (Fig 3B). No significant differences were detected between the ALS_short_ and ALS_long_ groups after FDR correction.

**Figure 3.**
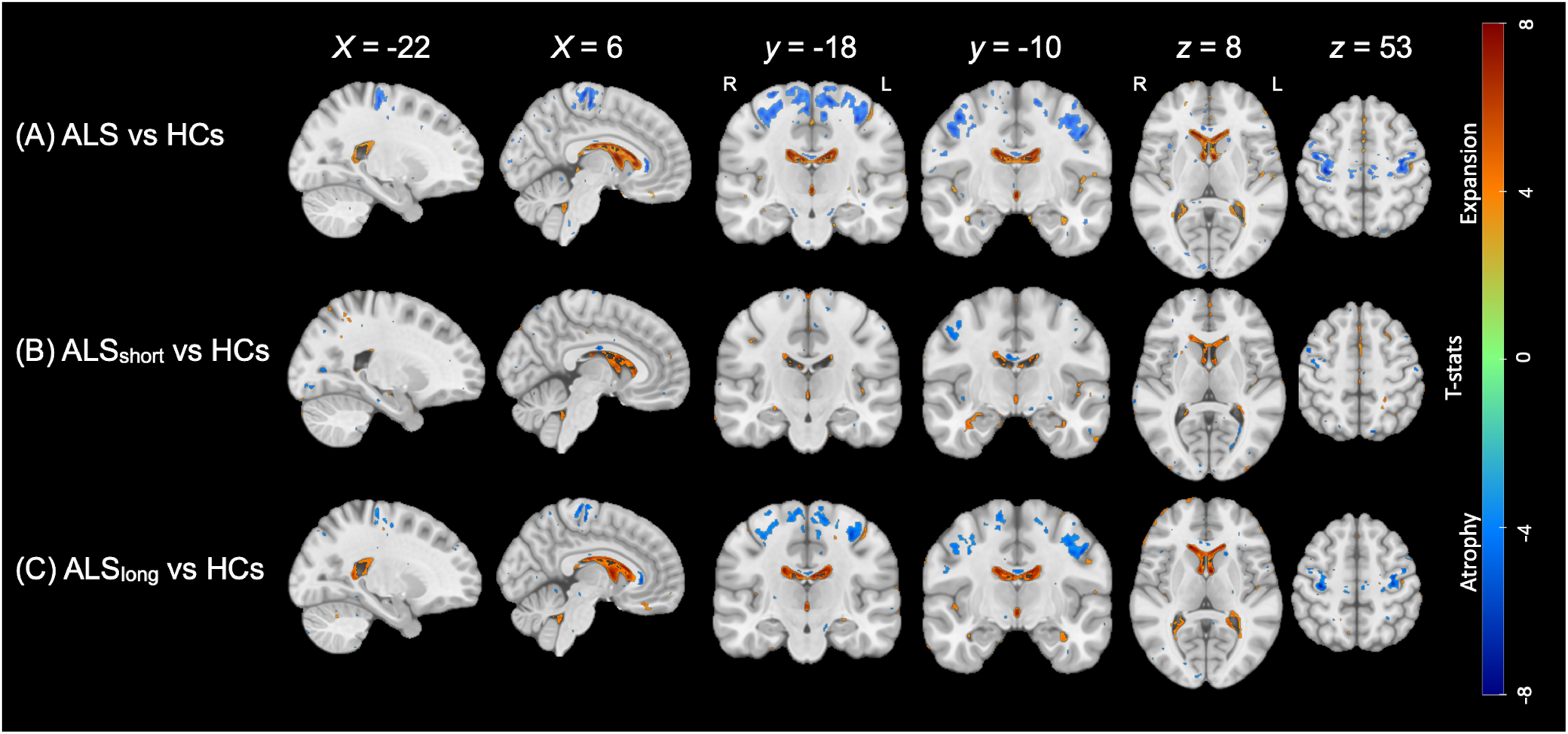
Longitudinal Volumetric Changes: T-statistic maps (p-FDR < 0.05) illustrate longitudinal differences (term Dx:ΔTime in the LME), overlaid on the MNI template. Rows display: ALS vs HCs **(A)** ALS_short_ vs HCs **(B)**, and ALS_long_ vs HCs **(C)**. ‘ALS_short_’ and ‘ALS_long_’ refer to patients who experienced the event prior to and after 24 months respectively. HCs represent healthy controls. Warmer colors highlight regions of enlargement (e.g., ventricular and sulcal regions), while cooler colors indicate areas of tissue atrophy.

### Association between brain atrophy and clinical symptoms

Our LME analysis revealed significant associations between regional atrophy and clinical manifestations of ALS (Fig 4A). Notably, CST atrophy was strongly linked to overall functional decline, as measured by ALSFRS-R and its subcategories (bulbar, motor, and respiratory), longer symptom duration, and faster disease progression. Precentral gyrus atrophy was associated with functional decline, bulbar and motor scores, reduced finger-tapping performance, prolonged symptom duration, and lower FVC. Brainstem atrophy was associated with lower ALSFRS-R scores, while paracentral atrophy was related to lower ALSFRS-R and motor scores, longer symptom duration, and poorer left foot tapping performance. Atrophy in the superior parietal region was also associated with lower ALSFRS-R and motor scores, and longer symptom duration. Thalamic atrophy was associated with lower ALSFRS bulbar scores, and ventral diencephalon atrophy showed associations with ALSFRS-R, bulbar scores, and longer symptom duration. Precuneus atrophy was linked to ALSFRS-R motor scores and impaired left foot tapping, while pars triangularis atrophy was associated with higher DPR and more advanced El Escorial diagnostic categories. Anterior thalamic radiation atrophy and expansions of the lateral and third ventricles were linked to longer symptom duration, with the third ventricle also associated with lower ALSFRS-R and motor scores. Finally, we identified critical factors related to time-to-event in uncensored ALS patients (Fig 4B). Shorter time-to-outcome was significantly associated with greater atrophy in the corpus callosum (t=3.84, *p*=0.0003) and CST (t=3.34, *p*=0.001), as well as lower ALSFRS-R scores (t=3.65, *p*=0.0005) and increased right-sided LMN burden (t=-3.22, *p*=0.002).

**Figure 4.**
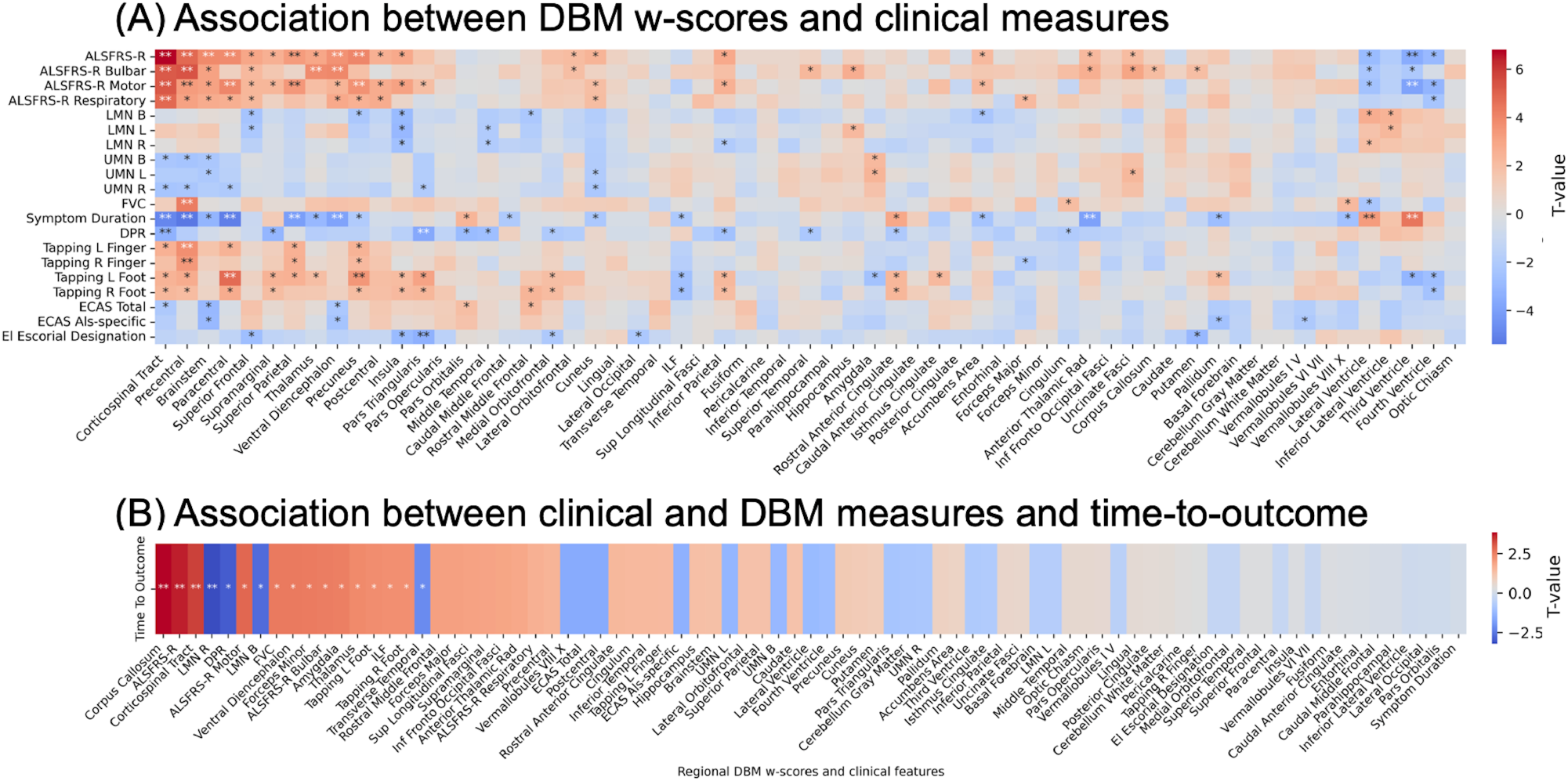
Association in measurements: **(A)** Associations between clinical measures (row) and regional DBM w-scores (columns) on longitudinal data. **(B)** Cross-sectional associations between uncensored patient’s time to outcome and all measurements. Analyses were corrected for age, sex and scanner effects. The color bar shows the T-value. Lower DBM w-scores indicate greater atrophy. *Significant associations at uncorrected *p* < 0.05. **Significant associations at FDR-corrected *p* < 0.05. ALSFRS-R: ALS Functional Rating Scale-Revised Total Score; ALSFRS-R Bulbar: sum of speech, salivation and swallowing scores; ALSFRS-R Motor: sum of fine and gross motor scores (writing, feeding, dressing, turning, walking and climbing); ALSFRS-R Respiratory: sum of dyspnea, orthopnea and respiratory insufficiency scores; FVC: Forced Vital Capacity; DPR: Disease Progression Rate; LMN: Lower motor neuron; ILF: Inferior Longitudinal Fasciculus.

### Survival prediction

Figure 5 illustrates examples of Individual Survival Curves (ISDs) predicted for uncensored patients from the test set of one cross-validation fold, using only clinical features (left) and a combination of regional DBM w-scores and clinical measurements (right). 50% survival probability is utilized for predicting event times in the 5-year observation window. In this example, incorporating imaging features led to an improvement in individual-level survival predictions as evident by a reduction in MAE-uncensored from 11 to 8.6 months.

**Figure 5.**
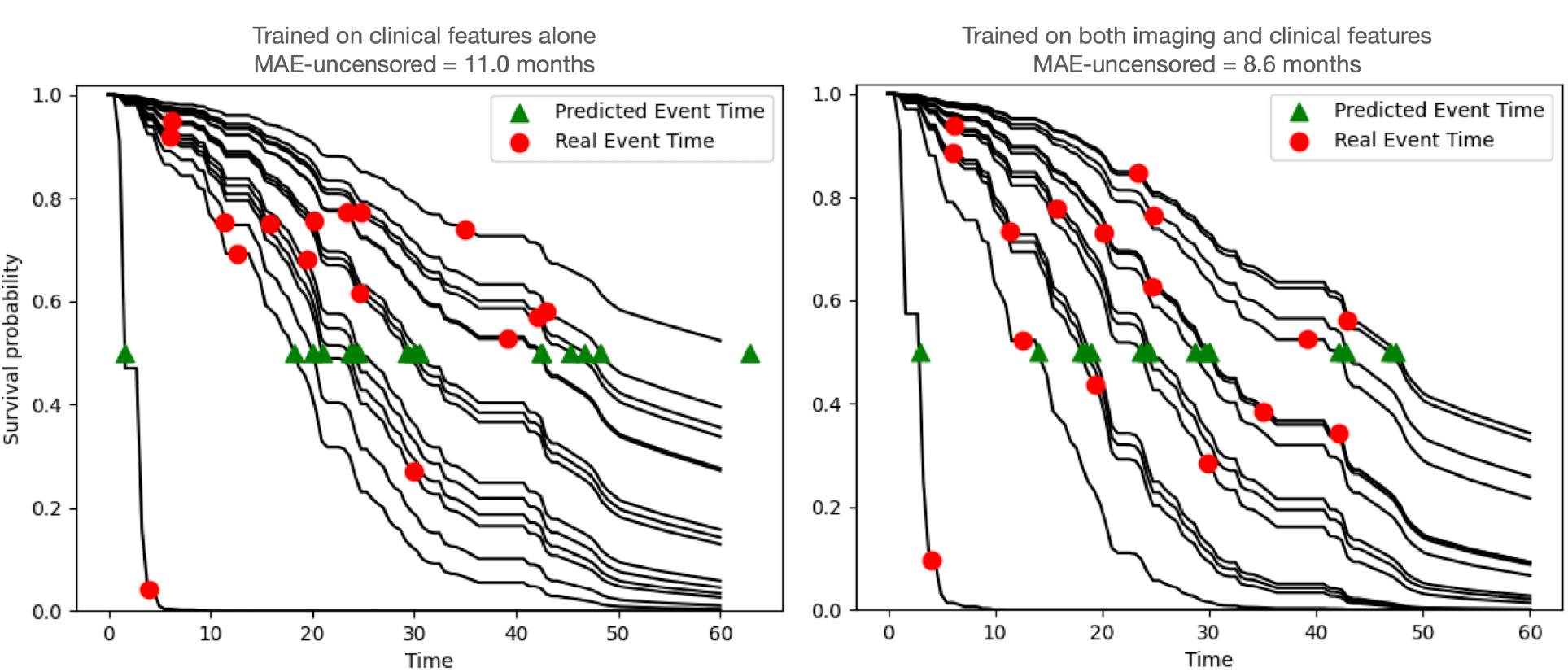
Individual Survival Distributions (ISDs) curves: Examples of Individual Survival Distribution curves predicted for uncensored and unseen patient data from one cross-validation fold, comparing the model trained solely on clinical features (left) versus the model trained on both imaging and clinical features (right). The individual’s predicted event time at 50% probability (green triangle) and the actual event time (red circle) are overlaid on the respective 5-year survival curves.

ISDs were predicted for all ALS patients through cross-validation, including all 87 censored patients. For the single-time prediction at *t=*24 months, 65 out of 87 censored patients were excluded as their censoring time occurred before 24 months, making it impossible to observe their status at the target time. Figure 6 presents the distribution of performance of the CPH model in terms of individual survival distribution (a) and binary classification predicting whether the time-to-event is prior to or after t=24 months (b). Evaluation is based on two sets of features: clinical features alone and combining clinical and DBM features. Adding imaging features to clinical data significantly improved model performance. The c-index distribution shows a clear shift towards higher values when DBM features are added, indicating superior discrimination between patients with different survival outcomes. Similarly, the IBS distribution demonstrates a shift towards lower values, suggesting improved overall prediction accuracy. Lower IBS signifies that predicted survival probabilities are closer to the true observed survival status of patients. Furthermore, both MAE-PO and MAE-Margin decrease when imaging features are incorporated, reflecting a reduction in prediction errors. Integrating DBM w-scores with clinical features enhanced survival prediction at 24 months, resulting in a shift toward higher balanced accuracy and discriminatory power, as evidenced by ROC-AUC. It also enhanced the model’s ability to correctly classify patients who experienced the event within (sensitivity) and after (specificity) 24 months. All distribution pairs were significantly different based on the Mann-Whitney test (*p*-values<0.05). Regardless of whether only clinical features or a combination of features was used, the models exhibited excellent calibration across the entire survival distribution in all iterations, as evidenced by D-calibration *p-* values>0.05. Training the model with only imaging features yielded lower performance compared to using clinical features alone (data not shown).

**Figure 6.**
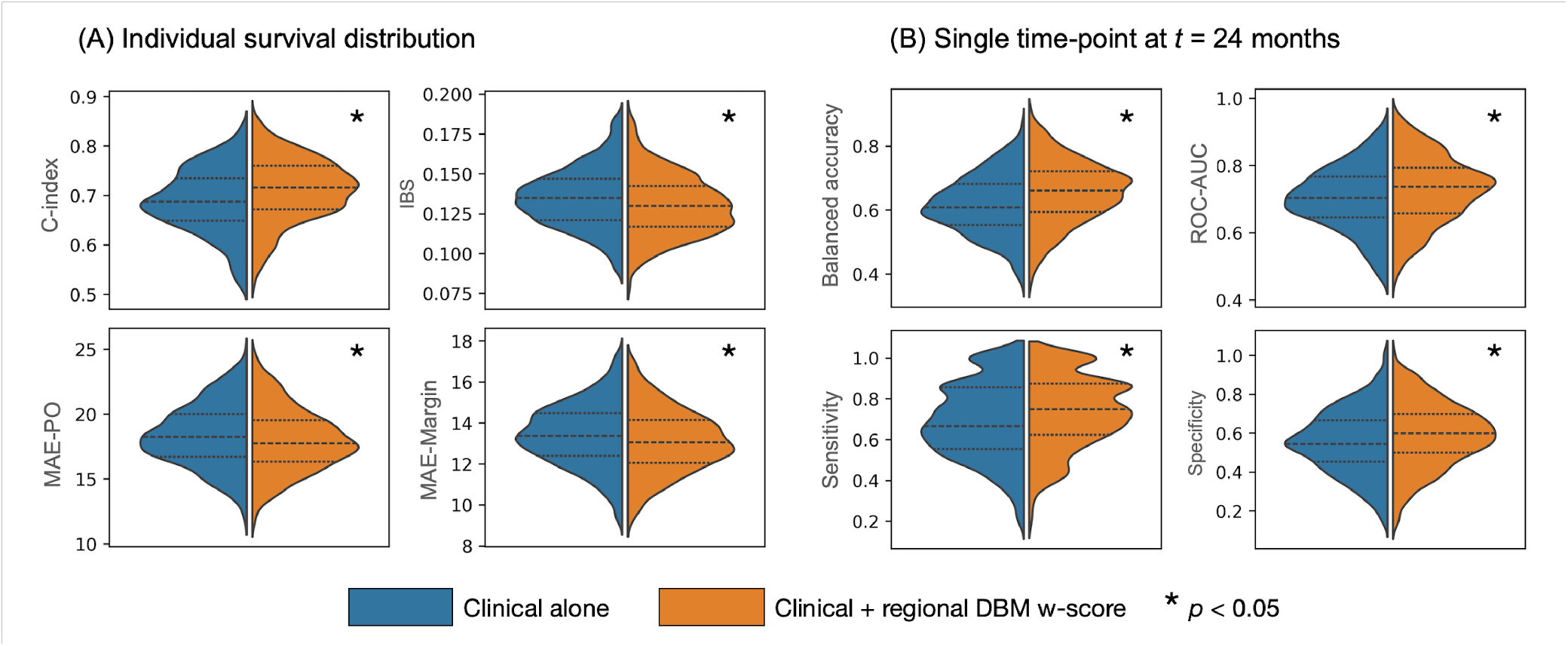
Survival prediction performance: Distribution of performance metrics across 100×5 cross-validation runs, comparing the use of clinical features alone (blue) to the inclusion of imaging features (orange). The dashed lines represent the median of each distribution, while the dashed-dot lines indicate the first (Q1) and third quartiles (Q3). Panel **(A)** illustrates the performance at the level of Individual survival distributions (ISDs). Panel **(B)** focuses on single-time point classification at t=24 months. The asterisk denotes a statistically significant difference, with a *p*-value less than 0.05. C-index: concordance index; IBS: integrated brier score; MAE: mean absolute error; MAE-PO: MAE using pseudo observations; ROC-AUC: mean area under the receiver operating characteristic curve.

### Feature importance as predictors of survival

The model trained on baseline clinical and imaging features identified the key predictors through nested cross-validation. The top 25 most important features are shown in Figure 7A. Among the significant clinical predictors were higher DPR, higher LMN burden in the right arm and leg, reduced scores in ALSFRS swallowing, salivation, handwriting, and total score, lower ECAS verbal fluency score, and decreased FVC. Additionally, atrophy in the corpus callosum, rostral middle frontal gyrus, thalamus, amygdala, and CST, was strongly associated with an elevated risk of the composite outcome. Panel (B) highlights GM regions where atrophy is linked to an increased risk of the event, while panel (C) illustrates WM tracts where atrophy is significantly associated with an elevated risk of the outcome.

**Figure 7.**
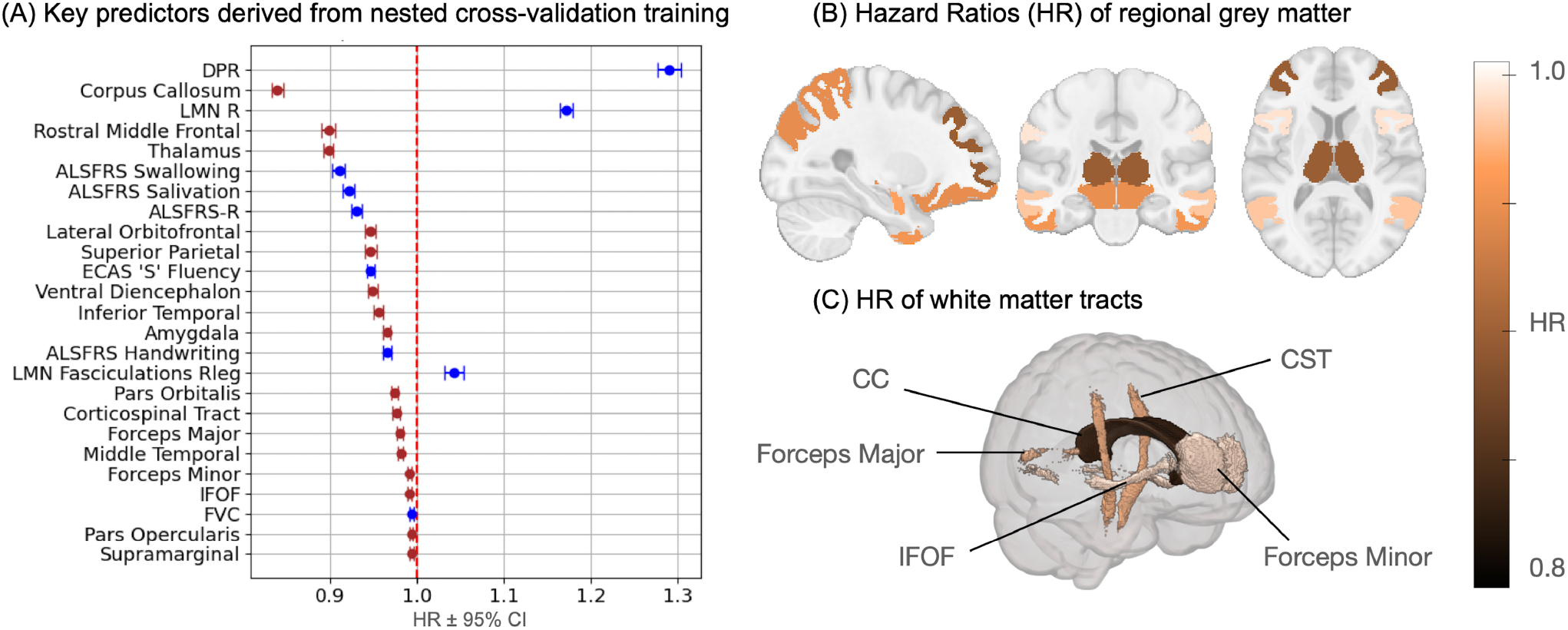
Key predictors: **(A)** Cross-sectional clinical and regional w-scores (DBM) features identified as predictors of survival outcomes by the CPH model during nested cross-validation training. Hazard ratio (HR) ± 95% confidence interval for clinical features are shown in blue, while those for regional DBM are shown in red. An HR value above 1 indicates that the increase in the feature’s value is associated with an increased risk of event, whereas an HR value below 1 indicates that a decrease in the feature’s value (for instance, atrophy in a brain region) is associated with an increased risk of event. **(B)** Sagittal, coronal, and axial views of significant GM regions from the CerebrA atlas^34^. **(C)** Brain glass visualizations showing significant WM tracts from the JHU50 atlas^35^ and the Allen atlas^36^ (specifically for the corpus callosum). **(B-C)** Key regions are colored according to their HR: darker colors (approaching HR=0.8) indicate that atrophy in these regions has a greater impact on survival, whereas regions tending toward lighter, white shades (HR=1) suggest a lesser influence. CST: corticospinal tract; CC: corpus callosum; IFOF: inferior fronto-occipital fasciculus.

## Discussion

This study aimed to investigate ALS disease-related cerebral atrophy patterns as quantified by DBM and whether incorporating DBM, alongside clinical features, improves the accuracy of models predicting individualized survival outcomes in ALS patients. Our key findings were as follows: 1. There were significant differences in the atrophy patterns in short versus long survival groups. 2. Both clinical features and regional brain atrophy were associated with survival. 3. Imaging combined with clinical features resulted in more accurate models than clinical features alone. 4. The strongest predictors of shorter survival were: atrophy in the corpus callosum, rostral middle frontal gyrus, and thalamus as well as higher DPR, greater LMN burden in the right arm and leg, and reduced scores on the ALSFRS. These findings highlight the potential of DBM as a valuable biomarker in ALS, offering a more granular and personalized approach to prognosis.

Our voxel-wise analyses revealed distinct atrophy patterns in ALS patients. Cross-sectionally, significant GM atrophy was observed in key motor-related regions, ventral diencephalon, amygdala, thalamus, and brainstem. Degeneration was also observed in WM, including the CST, superior longitudinal fasciculus, anterior thalamocortical tracts, and corpus callosum. These findings are consistent with the pattern of atrophy reported previously^9,37^. Moreover, an expansion of the third ventricle in individuals with ALS was noted. It is believed that an increased width of the third ventricle (WTV) serves as an indirect marker of subcortical brain atrophy, particularly affecting adjacent structures such as the thalamus. Previous studies have also reported a significantly larger WTV in ALS patients, reinforcing this association^46^.

Longitudinally, we observed progressive atrophy in somatomotor regions, corpus callosum, and superior longitudinal fasciculus, along with additional small areas of atrophy spread in GM and WM of the brain, reflecting the disease’s advancing nature over time. Further enlargement of ventricles and sulci was also observed. Comparing ALS patients who experienced disease events within or after 24 months revealed that the shorter survival group exhibited more pronounced atrophy at baseline. This suggests that early atrophy may be indicative of a more severe disease progression. Despite these observations, statistical differences between the two survival groups were relatively limited, with atrophy at baseline noted primarily in the body and splenium of the corpus callosum and the right-sided head of caudate. The limited statistical differences between the short and long survival groups could be attributed to the relatively low sample size and incomplete follow-up data – out of 178 ALS patients, only 101 could be classified as either short (41) or long (60) survivors. This constraint likely reduced the statistical power to detect subtle but potentially meaningful differences. Moreover, the higher attrition rate in the short survival ALS group may account for the fewer significant regions detected in the ALS short survival group compared to the long survival group. Specifically, the number of long survival ALS patients who completed visits 1, 2, and 3 were 60, 41, and 30, respectively, while the corresponding numbers for short survival ALS patients were 41, 18, and 9. This significant drop-off in participant numbers in the short survival group likely reduces the statistical power to detect longitudinal brain changes, making it more challenging to identify significant differences compared to the healthy controls and even more so compared to the long survival group.

The associations identified in our study underscore the extensive neurodegeneration observed in ALS, particularly highlighting the critical role of atrophy in the CST and precentral gyrus. These regions were strongly associated with overall functional decline, faster DPR, and both motor and bulbar impairments. Moreover, these findings indicate that ALS affects multiple facets of function beyond motor control, including bulbar and respiratory capabilities. Notably, atrophy in the thalamus and ventral diencephalon was linked to bulbar function, emphasizing their significance in ALS-related dysphagia and speech difficulties. The correlation between ventricular expansion and prolonged symptom duration suggests that ALS is characterized by widespread brain changes that extend beyond the primary motor pathways. Importantly, the consistent association between prolonged symptom duration and neurodegeneration highlights the need for early intervention strategies in ALS, which may improve patient outcomes. These results support the potential of using regional atrophy as a biomarker for monitoring ALS progression and guiding clinical interventions. Interestingly, while the corpus callosum is consistently recognized as a target in ALS, it did not correlate with any clinical measures. However, it did show a significant correlation with time-to-event and, more importantly, emerged as the most impactful imaging predictor identified by our survival model.

The integration of regional DBM features with clinical data resulted in a clear shift in model performance across key metrics. We observed notable improvements, including enhanced discrimination capacity, better calibration, reduced mean absolute error, and an increased ability to predict patient outcomes at the 24-month time point. While some overlap persists between the performance distributions with and without imaging features, indicating that the impact of DBM features may not be uniform across all cases, the overall trend toward superior performance is evident. Even when subtle, these enhancements carry significant clinical relevance, particularly in ALS, where even modest improvements in predictive accuracy can drive more personalized treatment strategies and potentially improve patient outcomes. This underscores the value of incorporating imaging biomarkers that capture disease-specific brain atrophy patterns, providing prognostic insights beyond what clinical data alone can reveal. These results are consistent with previous studies that have also demonstrated significant improvements in survival prediction by integrating imaging features with clinical data^12,14,15,47,48^.

Our survival model, which was trained on both imaging and clinical measures, identified several brain regions and clinical measures as key predictors associated with the outcome of death or respiratory failure in ALS patients. Notably, atrophy in the corpus callosum, rostral middle frontal, thalamus, amygdala, and CST significantly influenced the likelihood of these events. These findings highlight the critical contributions of motor, frontotemporal, subcortical, GM, and WM regions to survival outcomes in ALS. Moreover, these regions are consistent with those identified in earlier studies on ALS progression^12,13,16,49^. Additionally, critical clinical predictors identified by the model included the DPR, lower motor neuron burden on the right side, FVC, various components of the ALSFRS-R (including total score, swallowing, salivation, and handwriting) and verbal fluency on the ECAS. These findings emphasize the crucial role of motor, respiratory, and cognitive functions in predicting survival outcomes in ALS.

In a previous study employing ISD models^11^, only clinical features were selected, as cortical thickness from MR images did not enhance survival predictions. The difference in mean absolute error between clinical features alone and combined with imaging was not significant. In contrast, our study shows a clear improvement in prediction by incorporating DBM features from both grey and WM, highlighting the importance of WM atrophy, particularly in the corpus callosum and CST, which have been linked to ALS progression in previous studies^12,14^.

Traditional models provide a single-point survival prediction, missing ALS’s dynamic and heterogeneous nature. In contrast, ISD models predict survival probabilities over time, offering a more personalized prognosis crucial for ALS, given its variability in symptoms, progression, and treatment responses. Researchers and clinicians can use our model to produce individualized survival predictions for new ALS patients. For patients predicted to have faster disease progression, clinicians can apply more aggressive monitoring and therapeutic strategies. Additionally, similar to a previous approach to enrich Alzheimer’s disease clinical trials^50^, predicting individual decline can significantly enhance ALS clinical trials. By selecting participants who are more likely to show disease progression during the trial, researchers improve the study’s ability to discern the efficacy of treatments, focusing on participants who are most likely to benefit from the interventions.

Despite the promising results, several challenges need to be addressed to further enhance the robustness and generalizability of DBM-based prognostic models. One challenge is the inclusion of patients with diverse clinical and demographic characteristics. While the study included a relatively large and deeply phenotyped cohort, with an element of geographical diversity across multiple sites in Canada and the US, the generalizability of the findings to broader ALS populations remains to be validated on an independent dataset to ensure the models’ applicability across different clinical settings and populations. Moreover, while our models effectively account for censored patients —thereby avoiding the bias commonly introduced by excluding these patients^20,21^, our predictive accuracy could be further improved by reducing the current censoring rate, which stands at 52%. Increased survival follow-up could help address this issue and improve the robustness of our model’s predictions.

Advanced MRI techniques like DBM are transforming ALS biomarker development by quantifying regional atrophy in grey and WM, as well as ventricular and sulcal expansion, offering a detailed view of neurodegeneration. Our findings support integrating imaging with clinical data to better capture ALS’s complexity. Combining DBM with other imaging metrics could further enhance survival model accuracy. In conclusion, incorporating grey and WM imaging improves survival prediction and understanding of ALS progression. Continued advancements and collaboration are key to translating these insights into clinical practice, improving diagnosis, prognosis, and personalized care.

## Acknowledgments

We are grateful to the study participants, site PIs, research staff and MRI technologists. The authors also acknowledge use of Compute Canada (https://alliancecan.ca/en) resources for performing the image processing analyses in the presented work. This project was supported by research funds from a ALS-Canada Brain Canada discovery grant. Isabelle Lajoie is also supported by a postdoctoral fellowship from ALS-Canada and Brain Canada Foundation.

## Authors Contributions

I.L, S.K and M.D contributed to the conception and design of the study. I.L and M.D performed the analysis of the data. I.L drafted a significant portion of the manuscript and figures. I.L, S.K and M.D contributed significant intellectual content to the study and provided critical edits to the manuscript. The members of the Canadian ALS Neuroimaging Consortium (CALSNIC) performed the data acquisition. See supplementary materials Table S1 for a complete list of the CALSNIC group members.

## Data Availability Statement

Imaging and clinical data can be requested from the CALSNIC consortium (https://calsnic.org/)

