## supplementary materials for "Regional cerebral atrophy contributes to personalized survival prediction in ALS: a multicentre, machine learning, deformation based morphometry study"

### Supplemental Material

A. Exclusion criteria

Patients were included if they had signs of both UMN and LMN involvement upon initial diagnosis and met criteria for possible, probable, laboratory-supported probable or definite ALS according to the Revised El Escorial Criteria[23]. Patients aged below 40 years, those that received a diagnosis of other neurological or psychiatric conditions (e.g. bipolar disorder, brain trauma, epilepsy and depression), or patients with a baseline symptom duration (i.e., the time between symptom onset and the date of the first MRI scan) of more than five years, were excluded. Healthy control participants were excluded if they were aged below 40 years old or if they had history of cognitive impairment or neurological and psychiatric disorders. Participants who either did not undergo MRI scans or whose scans failed quality control assessments were also excluded.

B. Clinical evaluations

Clinical evaluation included the ALS Functional Rating Scale-Revised (ALSFRS-R)[24], from which the rate of disease progression rate (DPR) was estimated employing the formula: (48-ALSFRS-R)/symptom duration. Finger and foot tapping rates were calculated based on the average number of taps in 10 seconds over two independent trials from the left and right side. Left and right measurements were then averaged to obtain a unique score for finger and foot tapping. The Edinburgh Cognitive and Behavioural ALS Screen (ECAS), a multi-domain cognitive screening battery developed for patients with ALS[25] was also performed. Cognitive impairment was assessed using quantile regression-derived cutoffs for the North American version of the ECAS that accounts for age and education level[26]. A composite score was computed to evaluate the extent of UMN involvement by combining information from tone, muscle stretch reflexes, presence of Babinski sign and pseudobulbar affect[22]. Similarly, a single LMN burden composite score was measured by considering hyporeflexia, muscle atrophy, and presence of fasciculations. Additional clinical and demographic features of interest included site of (first symptom) onset, forced vital capacity (FVC), symptom duration at the first MRI visit, ethnicity, years of education, handedness, age and sex.

Supplementary Table 1: Membership of the Canadian ALS Neuroimaging Consortium (CALSNIC)

| Name | Title | Affiliation |
|---|---|---|
| Dr. Sanjay Kalra | Principal Investigator | University of Alberta, Edmonton, AB, Canada |
| Dr. Christopher [illegible] | Principal Investigator | University of Alberta, Edmonton, AB, Canada |
| Dr. Alan Wilman | Principal Investigator | University of Alberta, Edmonton, AB, Canada |
| Dr. Dean Eurich | Principal Investigator | University of Alberta, Edmonton, AB, Canada |
| Dr. Christian Beaulieu | Principal Investigator | University of Alberta, Edmonton, AB, Canada |
| Dr. Yee Hong Yang | Principal Investigator | University of Alberta, Edmonton, AB, Canada |
| Dr. Lawrence Korngut | Principal Investigator | University of Calgary, Calgary, AB, Canada |
| Dr. Richard Frayne | Principal Investigator | University of Calgary, Calgary, AB, Canada |
| Dr. Hannah Briemberg | Principal Investigator | University of British Columbia, Vancouver, BC, Canada |
| Dr. Lorne Zinman | Principal Investigator | University of Toronto, Toronto, ON, Canada |
| Dr. Simon Graham | Principal Investigator | University of Toronto, Toronto, ON, Canada |
| Dr. Angela Genge | Principal Investigator | McGill University, Montreal, QC, Canada |
| Dr. Annie Dionne | Principal Investigator | Université Laval, Quebec City, QC, Canada |
| Dr. Nicolas Dupré | Principal Investigator | Université Laval, Quebec City, QC, Canada |
| Dr. Christen Shoesmith | Principal Investigator | Western University, London, ON, Canada |
| Dr. Michael Benatar | Principal Investigator | University of Miami, Miami, FL, United States |
| Dr. Robert Welsh | Principal Investigator | University of Utah, Salt Lake City, UT, United States |
